# Increases but pronounced state-level disparities in semaglutide prescribing to Medicaid patients from 2018 to 2022

**DOI:** 10.1101/2025.07.31.25332171

**Authors:** Anjolina L. Hsiao, Jennifer S. Agwagom, Maria Y. Tian, Rhudjerry E. Arnet, Faith A. Garasich, Omar E. Camacho, Angelique L. Starke, Christina H. Jang, Zuleima I. Mero, Kenneth L. McCall, Brian J. Piper

## Abstract

Visual Abstract

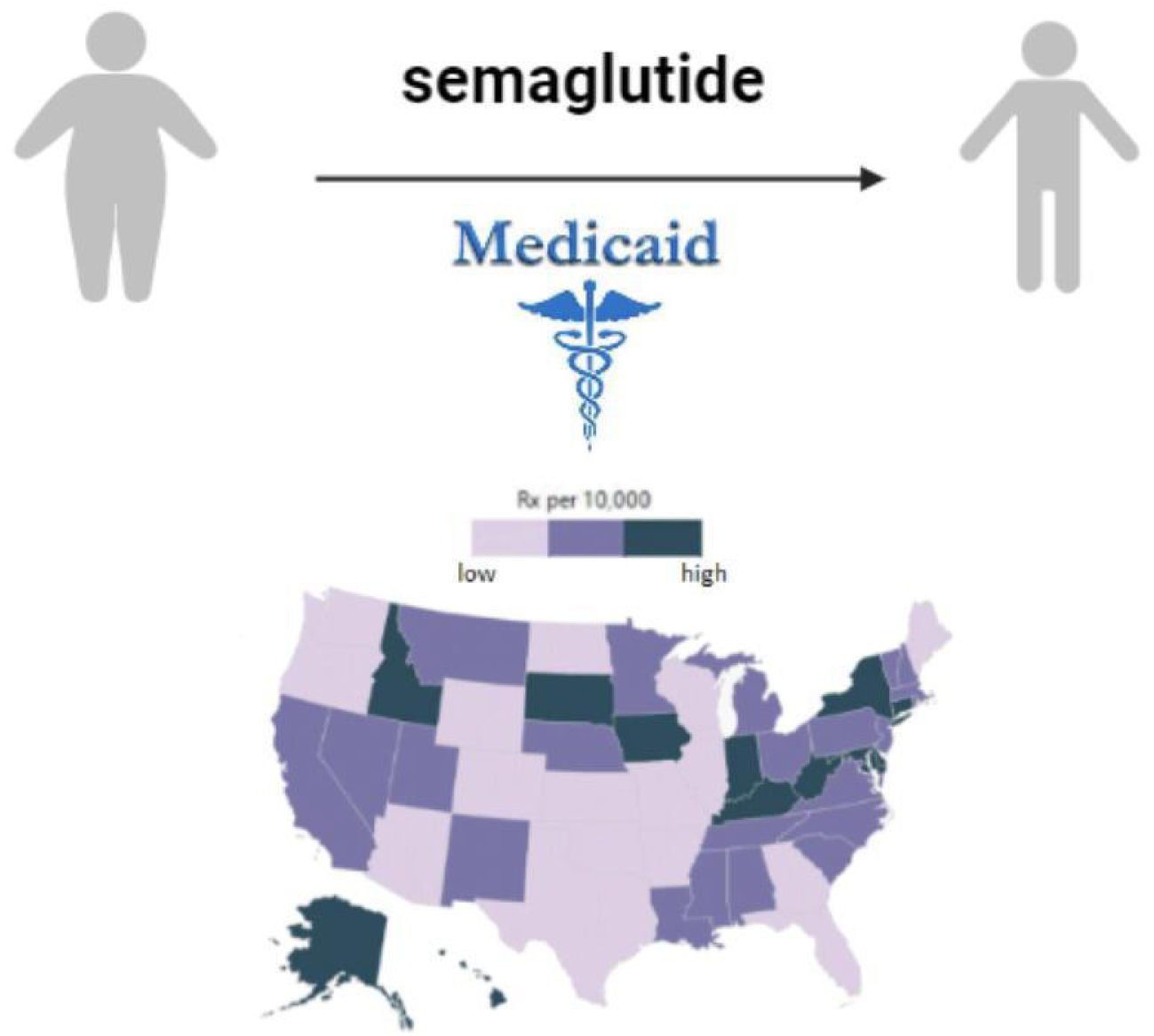

**Background:** Semaglutide is a glucagon-like peptide 1 receptor agonist that has been used to treat type 2 diabetes and for weight management since its approval in 2018 and 2021, respectively. Our research aimed to examine the geographic distribution of semaglutide prescriptions across the United States Medicaid program and determine the accessibility of this efficacious but costly to those on Medicaid.

**Methods:** Medicaid data on state drug utilization was collected to identify the number of semaglutide prescriptions distributed in each state per quarter from 2018 to 2022 and standardized them per 10,000 Medicaid enrollees. States whose rates fell outside the 95% confidence interval were considered significant outliers. Preferred drug lists for 2019-2022 were retrieved from state Medicaid website archives to explore coverage policies.

**Results:** National semaglutide prescribing in Medicaid rose steadily from 2018 to 2022. Notably, states in the Midwest, West, and Northeast regions showed the largest increases. After adjusted for enrollment, Indiana led usage of semaglutide from 2018 to 2020 at 185.6 prescriptions/10,000 enrollees. A notable surge in semaglutide prescriptions was observed in West Virginia in 2021. Conversely, states within the South and lower Midwest regions recorded the lowest prescription rates, with Arkansas (0.1 prescriptions/10,000 enrollees) ranking the lowest in 2020. Prescription volume did not correlate with obesity prevalence, per enrollee Medicaid spending, or the percentage of non-White population.

**Conclusion:** Semaglutide use within Medicaid climbed rapidly, but unevenly. Marked state-level variations highlight potential barriers to access despite the drug’s growing clinical importance.

## 1. Introduction

Heart disease and diabetes stand as the leading cause of mortality within the United States and are both associated with the prevalence of obesity.^1,2^ Semaglutide, a glucagon-like peptide-1 receptor agonist (GLP-1 RA), has gained approval for the treatment of both type 2 diabetes and obesity.^3^ Its mechanism of action, targeting discrete brain structures governing appetite and food consumption, renders it particularly efficacious.^3^ Semaglutide decreased preference for high-fat foods, as well as stimulated the production of insulin and inhibited the secretion of glucagon to facilitate the reduction of glucose absorption.^3,4^ Semaglutide gained FDA approval for injectable formulations in 2017 for type 2 diabetes management, followed by an oral dosage in 2019.^5^ The subsequent approval of a second injectable formulation in 2021 aimed at obesity further emphasizes its therapeutic versatility.

While injectable semaglutide has garnered approval for weight loss management, oral semaglutide requires a diagnosis of type II diabetes for coverage, highlighting potential barriers to access and warranting further investigation into semaglutide’s use across Medicaid. The high cost of semaglutide presents another important challenge. The average cost of a single semaglutide treatment is approximately $1561, with variability from $970 to $2,120 in the absence of insurance coverage.^6^

Access to semaglutide through Medicaid may vary considerably across states due to differences in preferred drug lists (PDLs). The status of preferred versus non-preferred formulations of the drug varies by state and their recommendations for when the drug can gain authorization. PDLs determine whether patients have coverage for certain medications through their insurance.Although no studies have specifically examined semaglutide’s inclusion on Medicaid PDLs, these state-level policy differences likely create substantial disparities in patient access. Given the substantial burden of type 2 diabetes and obesity within the US and the potential for unequal access to this effective but expensive therapy, we sought to explore geographic patterns in semaglutide prescribing for those with Medicaid coverage and identify disparities in access across states.

## 2. Materials Methods

### 2.1 Procedures

Data was obtained from the Medicaid Drug Utilization Database^7^ from 2018 to 2022 by quarter. Preferred drug lists for 2019-2022 were found using systematic search of each state’s Medicaid archives and frequent scouring of archives through Google. Information regarding PDLs was found utilizing each state’s Medicaid archive to locate mid-year drug lists for years 2018 through 2021. States without proper archives were supplemented with searches of Medicaid preferred drug lists in google and through use of the internet archive ‘Wayback Machine’^8^. Metformin was also obtained as a comparison substance. Procedures were approved as exempt by the Geisinger IRB.

### 2.2 Data-analysis

The number of prescriptions per 10,000 Medicaid enrollees was calculated. Scatterplots were used to determine the correlation between percent obese, percent non-white population, and dollars spent per Medicaid enrollee with the prescriptions per 10,000 Medicaid enrollees. GraphPad (v9.4.1, San Diego, California) aided in the construction of the scatter plots and JMP (v17, Cary, North Carolina) created heatmap.

Statistical analysis was performed by first calculating the national mean of the standardized prescription rate from 2018 to 2022. The 95% confidence interval was then found to determine states that were statistically elevated from the national mean. The fold difference was calculated between the states with the highest and lowest non-zero prescriptions per 10,000 enrollees.

## 3. Results

Figure 1 illustrates a marked increase in injectable prescriptions beginning in 2018 compared to an increase in oral prescription during 2019. This substantial rise in both oral and injectable formulations over two different time periods demonstrates the escalating adoption of semaglutide. Notably, Indiana maintained its position as the state with the highest prescribing rate in both 2018 and 2019, as shown in Supplemental Figure B and Supplemental Figure C, respectively. From 2018 to 2020, Indiana increased its prescription rate by over 13-fold.

**Figure 1:**
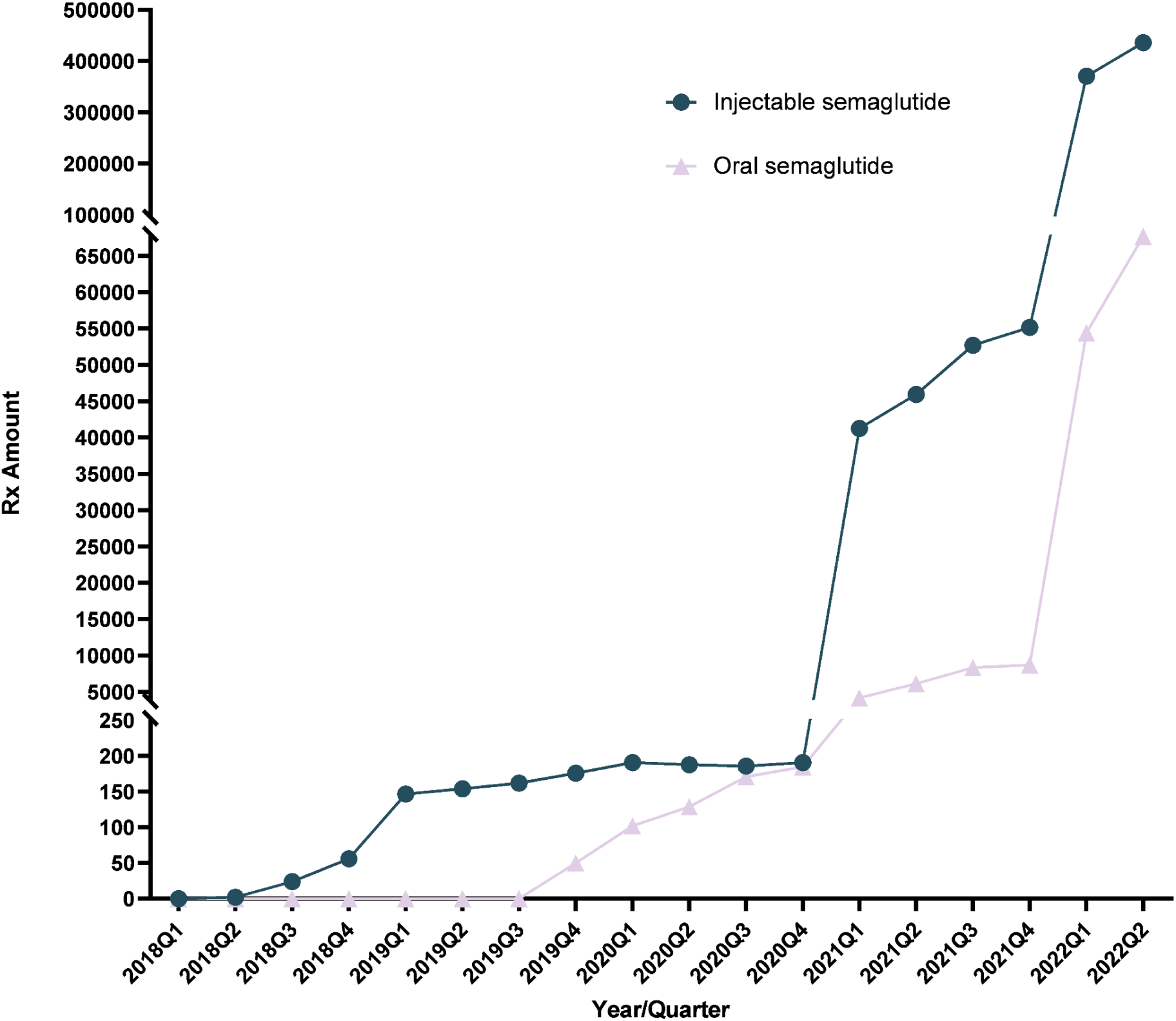
Medicaid prescriptions per quarter for oral and injectable semaglutide from 2018 to 2022.

Conversely, Arkansas persists as the state with the most restrained prescription rate, revealing minimal increases from 2018 through 2022. Moreover, West Virginia has raised its semaglutide prescription rate by over 200-fold from 2018 to 2021, emerging as the leading state in terms of prescriptions in 2021 (272.8/10,000 enrollees).

Upon calculating the prescriptions in each state per 10,000 Medicaid enrollees, there was a disparity between each state. Figure 3 shows Indiana having the highest rate of semaglutide prescriptions (185.6), compared against Arkansas, which registers the lowest rate (0.1) for 2020. Indiana, Iowa, Hawaii, and West Virginia exceeded the 95% confidence interval (lower end = 133.4), making them statistically significant. The discrepancy between the states with the highest and lowest prescription per enrollee ratio is staggering, amounting to an 1,856-fold difference between Indiana (185.6) and Arkansas (0.1). Between quarter 4 of 2020 and quarter 1 of 2021, injectable semaglutide prescriptions rose by over 200-fold, while oral semaglutide prescriptions witnessed a surge between quarter 4 of 2021 and quarter 1 of 2022, escalating by a factor of 6.

**Figure 2:**
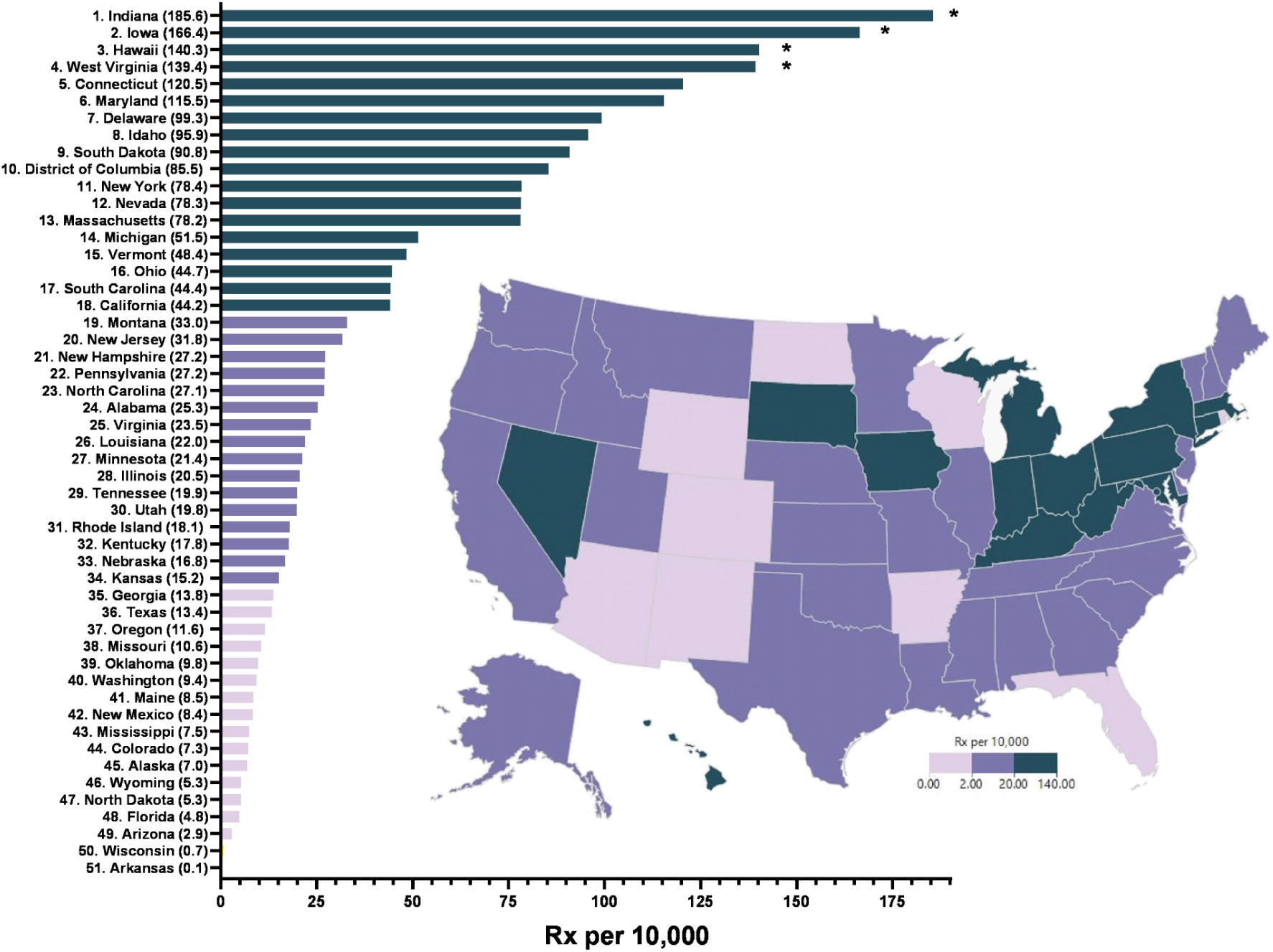
Heatmap and ranking of semaglutide (oral and injectable) prescriptions per 10,000 Medicaid Enrollees in 2020. Asterisk represents states that were significantly elevated relative to the national mean (Mean = **43.1** prescriptions, SD = **46.0**).

**Figure 3:**
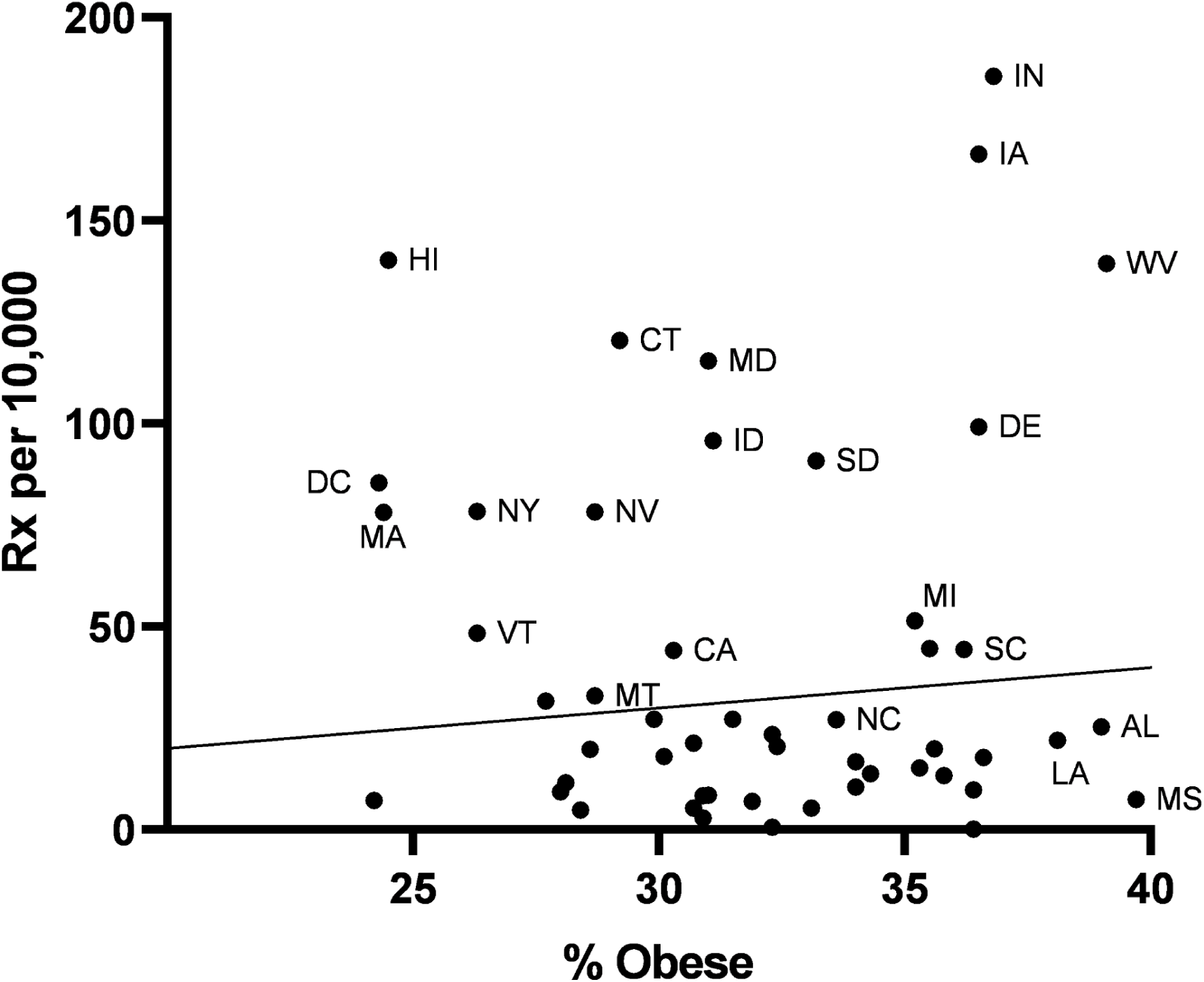
Scatterplot illustrating the non-significant association (r(49) = 0.017. p > .05) of percent obesity according to the Behavior Risk Factor Surveillance System by semaglutide prescriptions (oral and injectable) per 10,000 Medicaid enrollees in 2020.

Supplemental Figure D reveals Connecticut as the top prescribing state (278.5) during the first two quarters of 2022. The highest amount of both oral and injectable semaglutide prescribed was during the second quarter of 2022, with 67,690 and 418,868 prescriptions respectively.

Figure 3 shows no association between semaglutide prescribing and obesity (r(49) = 0.017, p > 0.05). Likewise, there was no correlation between dollars spent per enrollee and prescription rate (r(49) = 0.210, p > 0.05), as shown in Supplemental Figure F. Similarly, state demographics (percent non-White) are not associated with prescribing (r(49) = 0.012, p > 0.05), seen in Supplemental Figure G.

## 4. Discussion

Semaglutide prescriptions among Medicaid patients increased markedly from 2018-2022, showing regional disparities across prescribing states. Research on semaglutide prescribing among private insurance patients showed similar patterns, with semaglutide prescriptions increasing markedly around quarter 4 of 2020.^7^ Substantial increases occurred again in quarter 4 of 2021 and quarter 1 of 2022, with semaglutide prescribing among private insurance patients skyrocketing in the same quarters.^9^

State disparities in semaglutide prescriptions revealed that in particular, states within the South and Midwest had lower semaglutide prescription rates with Arkansas, Wisconsin, Wyoming, and North Dakota having the lowest prescribing rates. On the other hand, the Northeast and Northwest had higher semaglutide prescription rates with Indiana and Hawaii consistently having the highest prescribing rates. The state level disparity between Indiana, the highest prescribing state, and Arkansas, the lowest prescribing state was remarkable with an 1,856-fold difference in prescription per enrollee in 2020. Similar research on semaglutide prescribing across all payment options indicated similar elevations with the biggest increase happening in the end of 2020 to the beginning of 2021 and again in the beginning of 2022, consistent with our findings.^10^ This is contrasted with our findings of metformin with a difference of six fold in prescription per enrollee in 2020 (Supplemental Figure I), highlighting the pronounced state level disparities displayed in semaglutide prescribing among Medicaid enrollees.

Obesity prevalence, Medicaid expenditure per enrollee, preferred drug lists, and percent non-White population had no correlation with state level disparities in prescription rates. Prior research has found racial disparities showing that the percentage of adults eligible for semaglutide was highest in Black adults followed by Hispanic adults, but their access to semaglutide was limited due financial barriers.^11^ Despite these findings, our investigation shows that there was no correlation between injectable and oral semaglutide formulations per 10,000 Medicaid enrollees and the percent non-White population in each state. The difference in findings suggests a need for research exploring the accessibility and utilization of diabetes medication among different non-white populations enrolled in Medicaid programs.

Limitations of this study include a lack of focus on individual level variables including age of Medicaid enrollees as well as indications for use of semaglutide across brand name formulations. Future studies should consider these factors as a possible explanation for state level disparities in semaglutide prescribing and how state dependent coverage restrictions further influence the prescribing rate. Future investigations are needed to explore use of other GLP-1 agonists as semaglutide is one of six agents in this drug class. The pandemic is also likely to have affected the increased prescription rate of semaglutide but needs to be investigated further. The recency of semaglutide’s approval by the FDA for type 2 diabetes and obesity minimizes the information currently available.

## 5. Conclusion

This study identified appreciable increases in semaglutide prescribing nationally but also pronounced state level disparities to Medicaid patients. While some states demonstrated robust uptake of this effective therapy, others lagged significantly, creating inequitable access patterns across the US. Further investigation is warranted to determine the underlying causes of these state level differences and to develop targeted interventions that ensure equitable access to GLP-1 agonists. Addressing these disparities would maximize the potential to combat the US obesity epidemic.

## Supporting information

Raw Data

## Data Availability

All data produced are available at Medicaid.gov. All sorted data sets will be made available with the manuscript.

## Acknowledgements

This research was supported by HRSA (D34HP31025). Thank you to Maureen Murtha, Lavinia Harrison, MS, and Bill Rigotti for support.

ALH was affiliated with Thomas Jefferson University and Geisinger College of Health Sciences at the time of the study and is currently affiliated with Lake Erie College of Osteopathic Medicine. ALS was affiliated with Temple University at the time of the study and is currently affiliated with Miller School of Medicine at University of Miami.

## Disclosures

MT and BJP are supported by the Geisinger Academic Clinical Research Center. The other authors have no disclosures.

## Supplemental Figures

**Supplemental Figure A:**
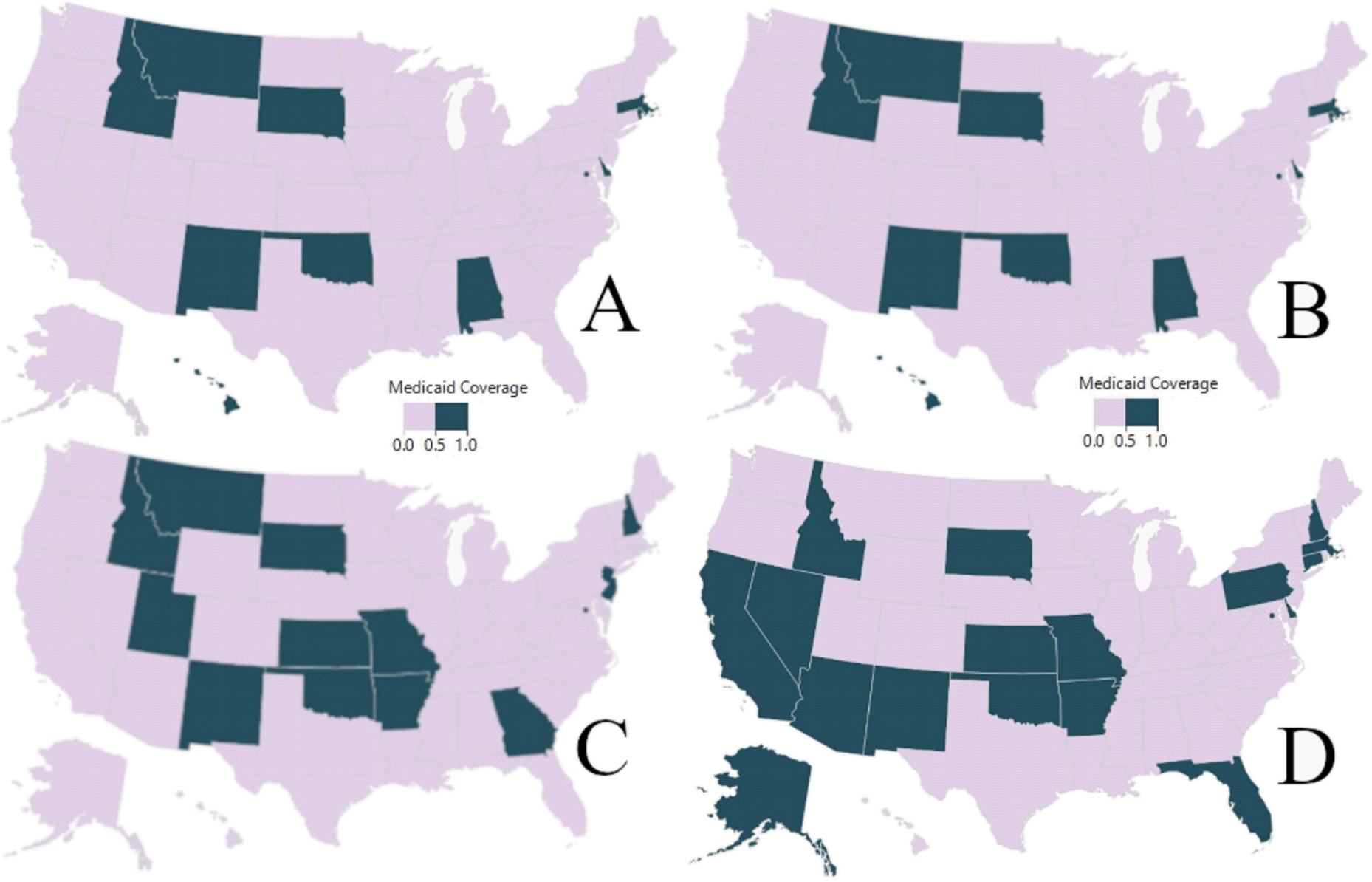
**(A)** Heatmap of states with semaglutide (oral and injectable) on preferred drug list for Medicaid in 2018. **(B)** Heatmap of states with semaglutide (oral and injectable) on preferred drug list for Medicaid in 2019. **(C)** Heatmap of states with semaglutide (oral and injectable) on preferred drug list for Medicaid in 2020. **(D)** Heatmap of states with semaglutide (oral and injectable) on preferred drug list for Medicaid in 2021.

**Supplemental Figure B:**
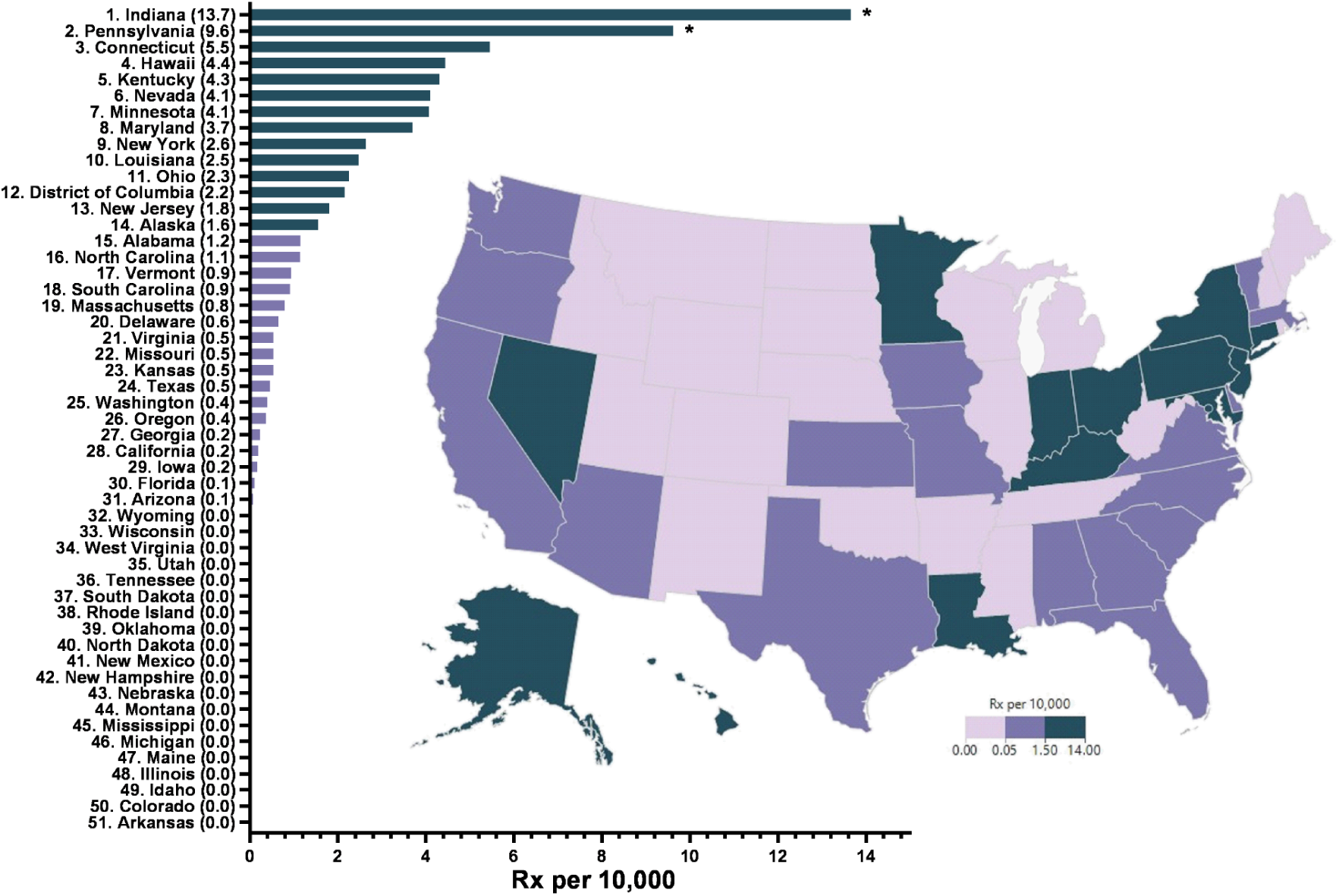
Heatmap and ranking of semaglutide (oral and injectable) prescriptions per 10,000 Medicaid Enrollees in 2018. Asterisk represents states that were significantly elevated relative to the national mean (Mean = 1.4 prescriptions, SD = 2.5).

**Supplemental Figure C:**
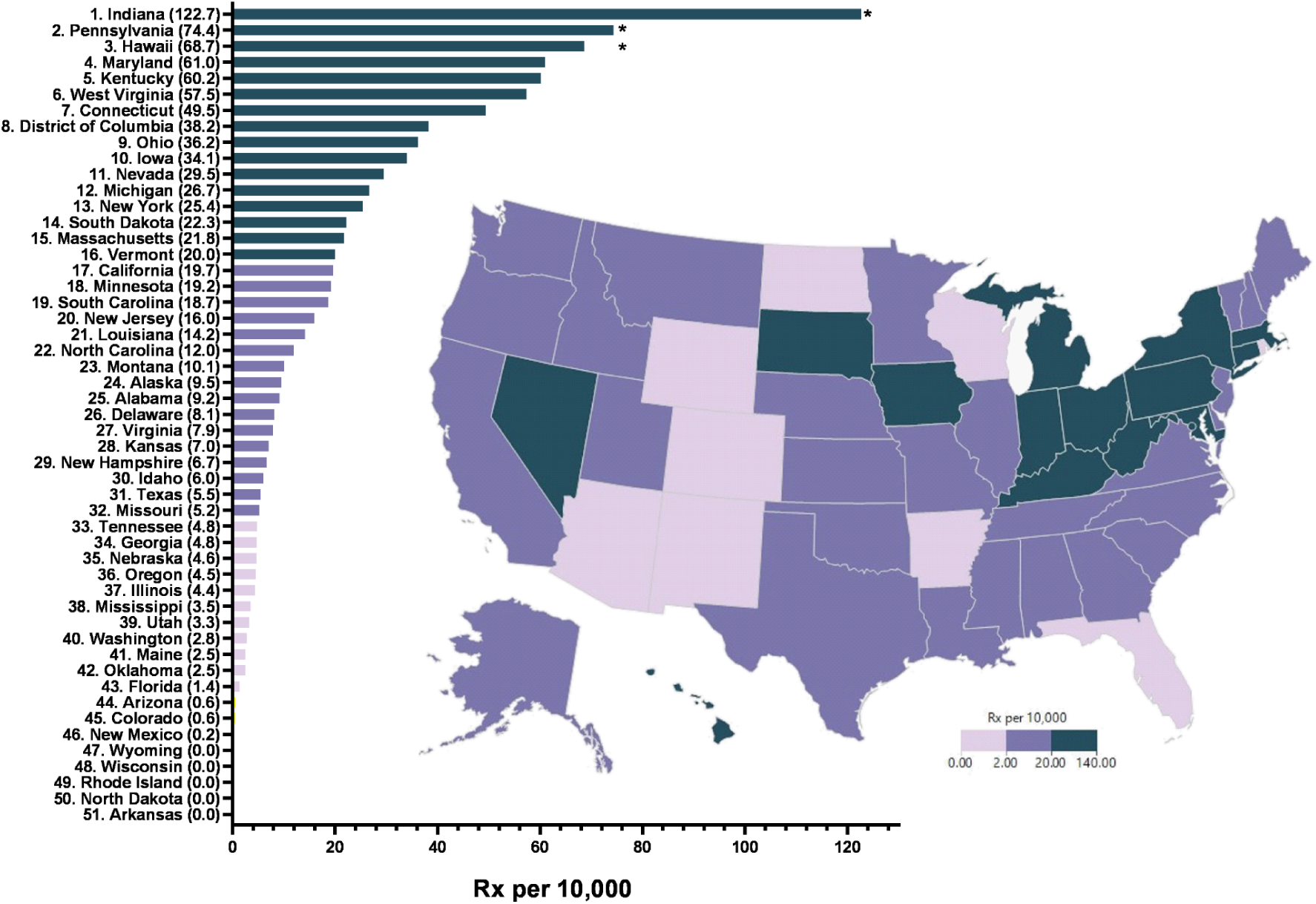
Heatmap and ranking of semaglutide (oral and injectable) prescriptions per 10,000 Medicaid Enrollees in 2019. Asterisk represents states that were significantly elevated relative to the national mean (Mean = **18.9**, SD = **24.3**).

**Supplemental Figure D:**
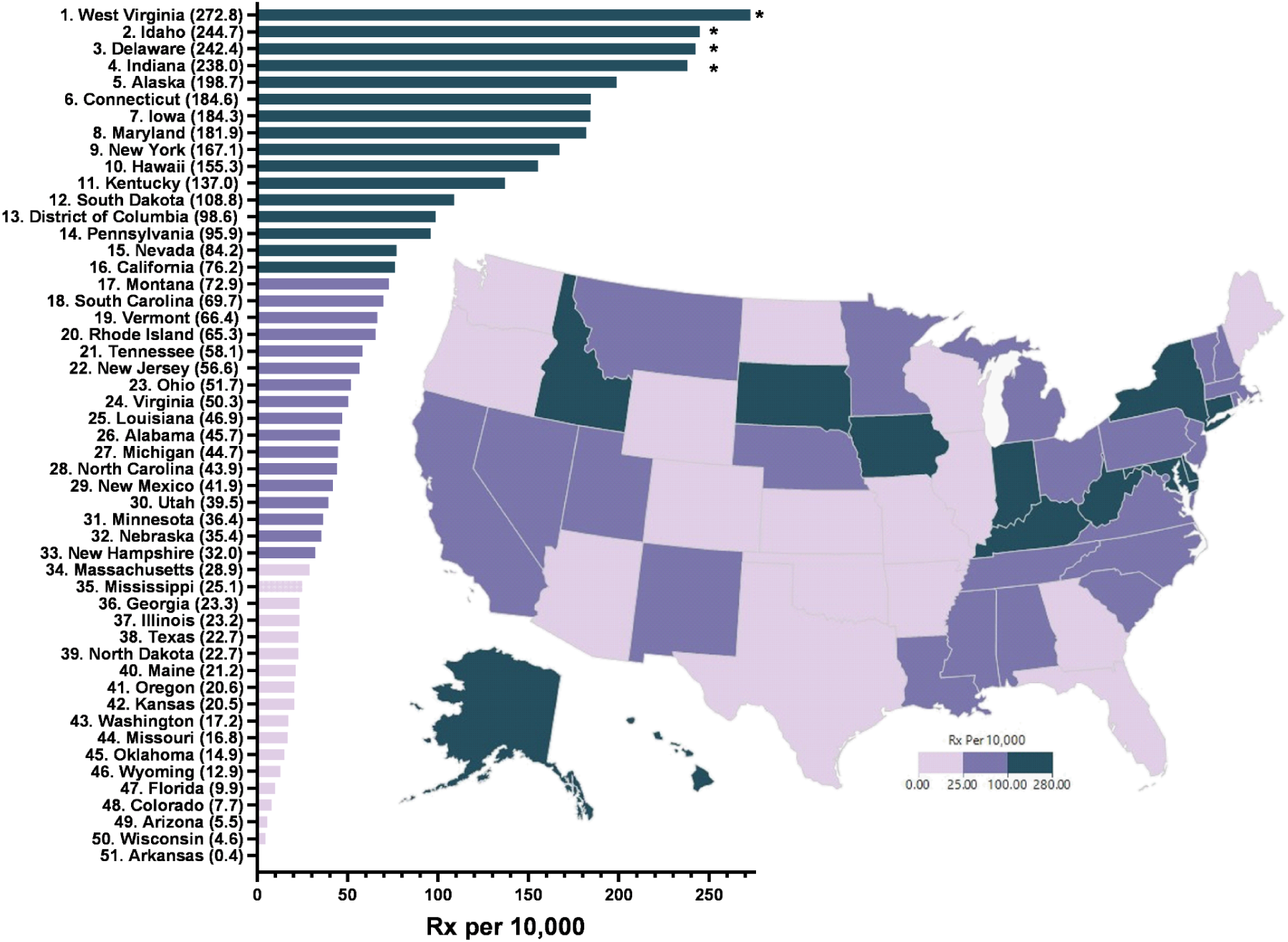
Heatmap and ranking of semaglutide (oral and injectable) prescriptions per 10,000 Medicaid Enrollees in 2021. Asterisk represents states that were significantly elevated relative to the national mean (Mean = **75.0** prescriptions, SD = **72.7**).

**Supplemental Figure E:**
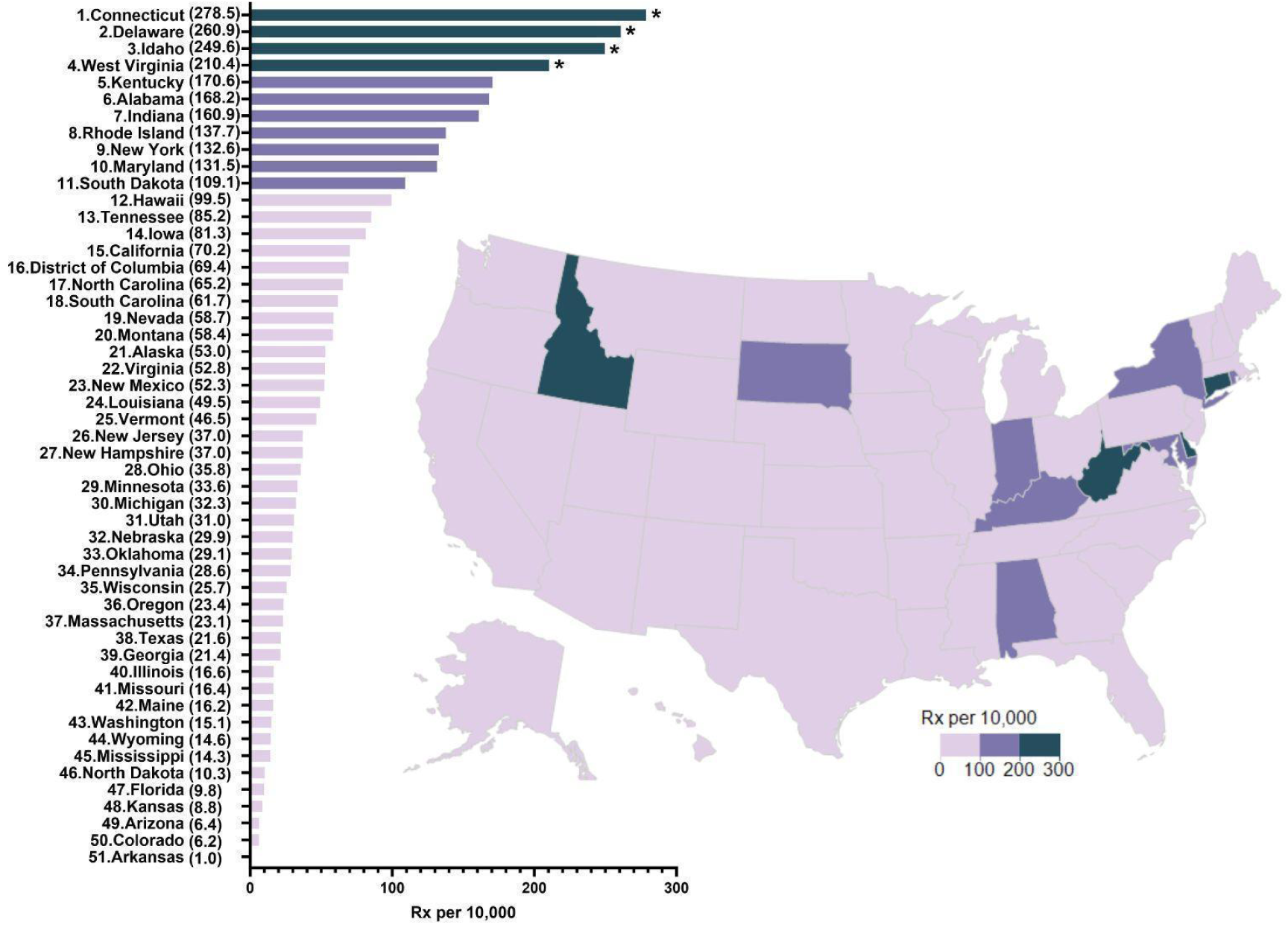
Heatmap and ranking of semaglutide (oral and injectable) prescriptions per 10,000 Medicaid Enrollees in Q1 and Q2 of 2022. Asterisk represents states that were significantly elevated relative to the national mean (Mean = **67.8** prescriptions, SD = **69.6**).

**Supplemental Figure F:**
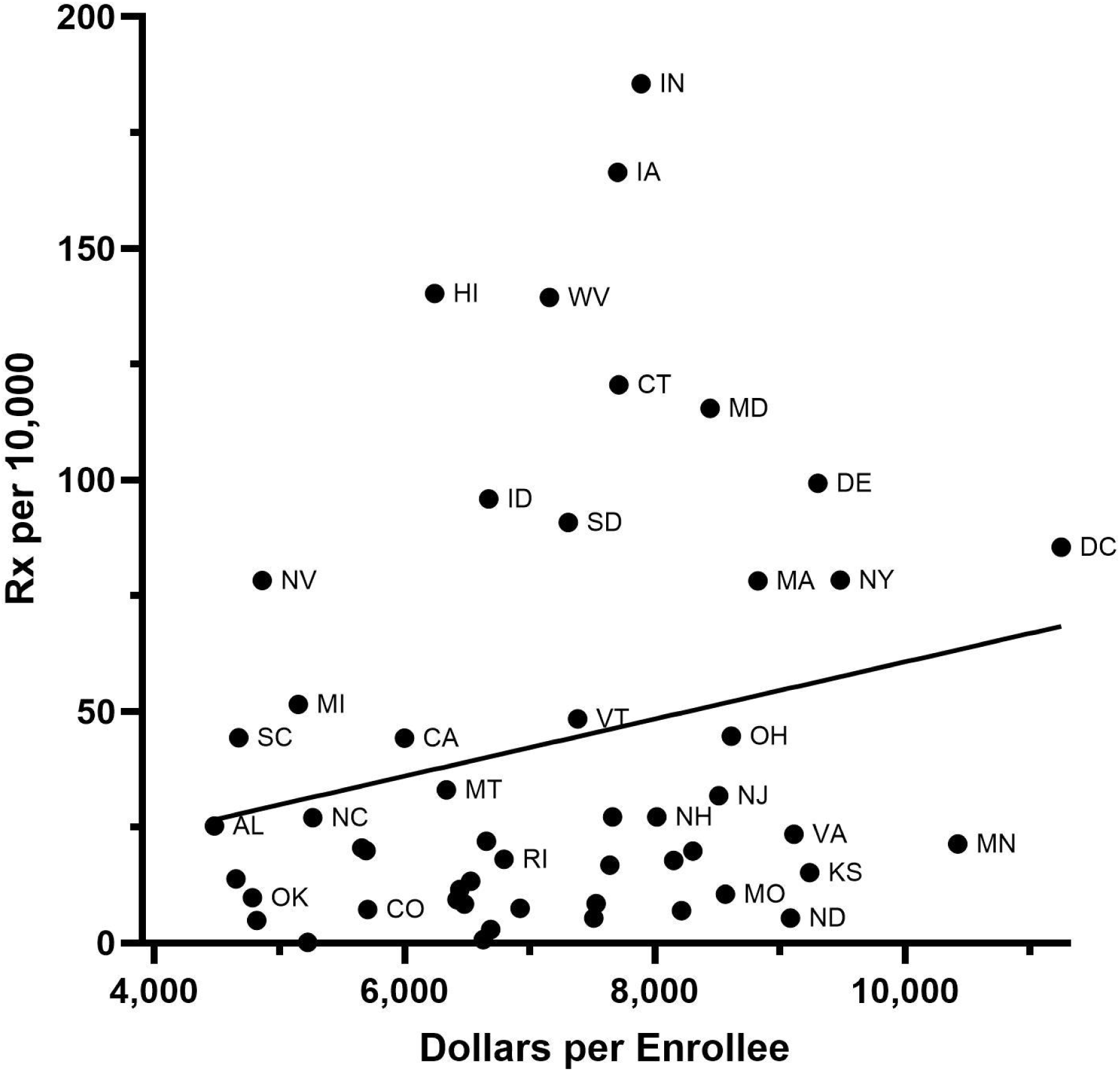
Scatterplot illustrating the non-significant association (r(49) = 0.210, p > .05) between injectable semaglutide prescriptions per 10,000 Medicaid enrollees and the Medicaid spending per enrollee in each state in 2020.

**Supplemental Figure G:**
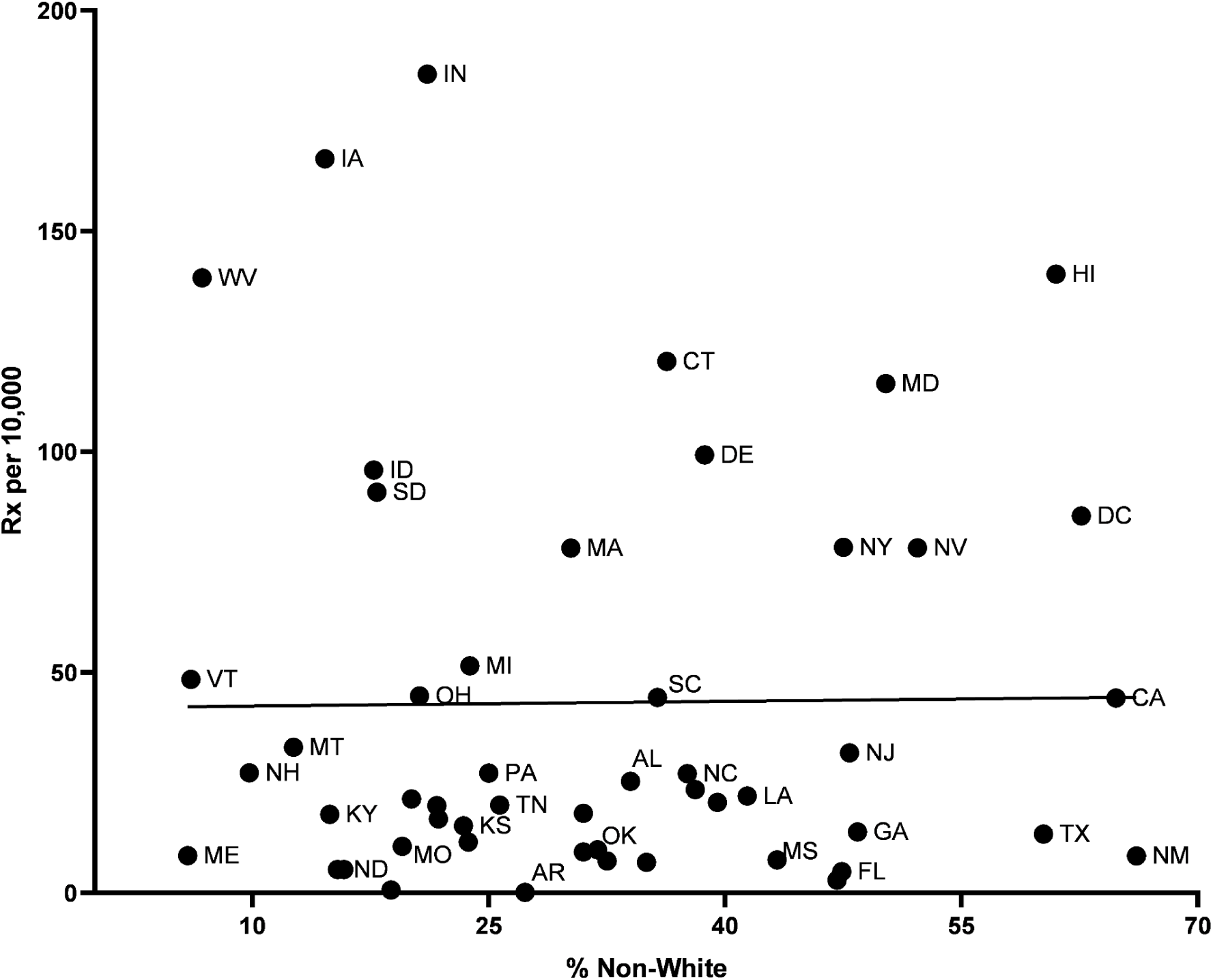
Scatterplot illustrating the non-significant association (r(49) = XY. p > .05) between injectable and oral semaglutide prescriptions per 10,000 Medicaid enrollees and the percent non-white population in each state in 2020.

**Supplement Figure H:**
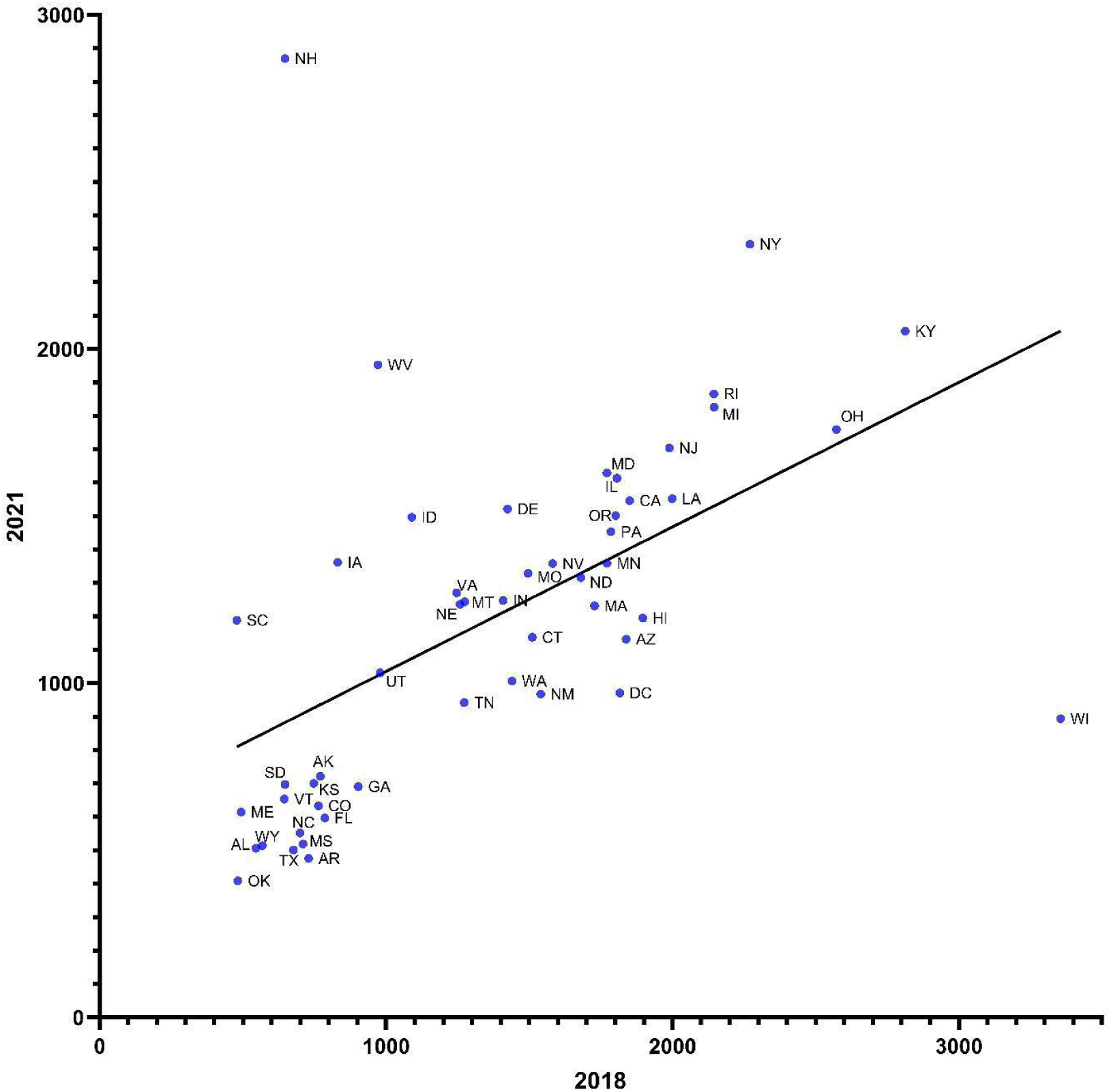
2018 vs 2021 state data on Rx per 10,000 enrollees

**Supplemental Figure I:**
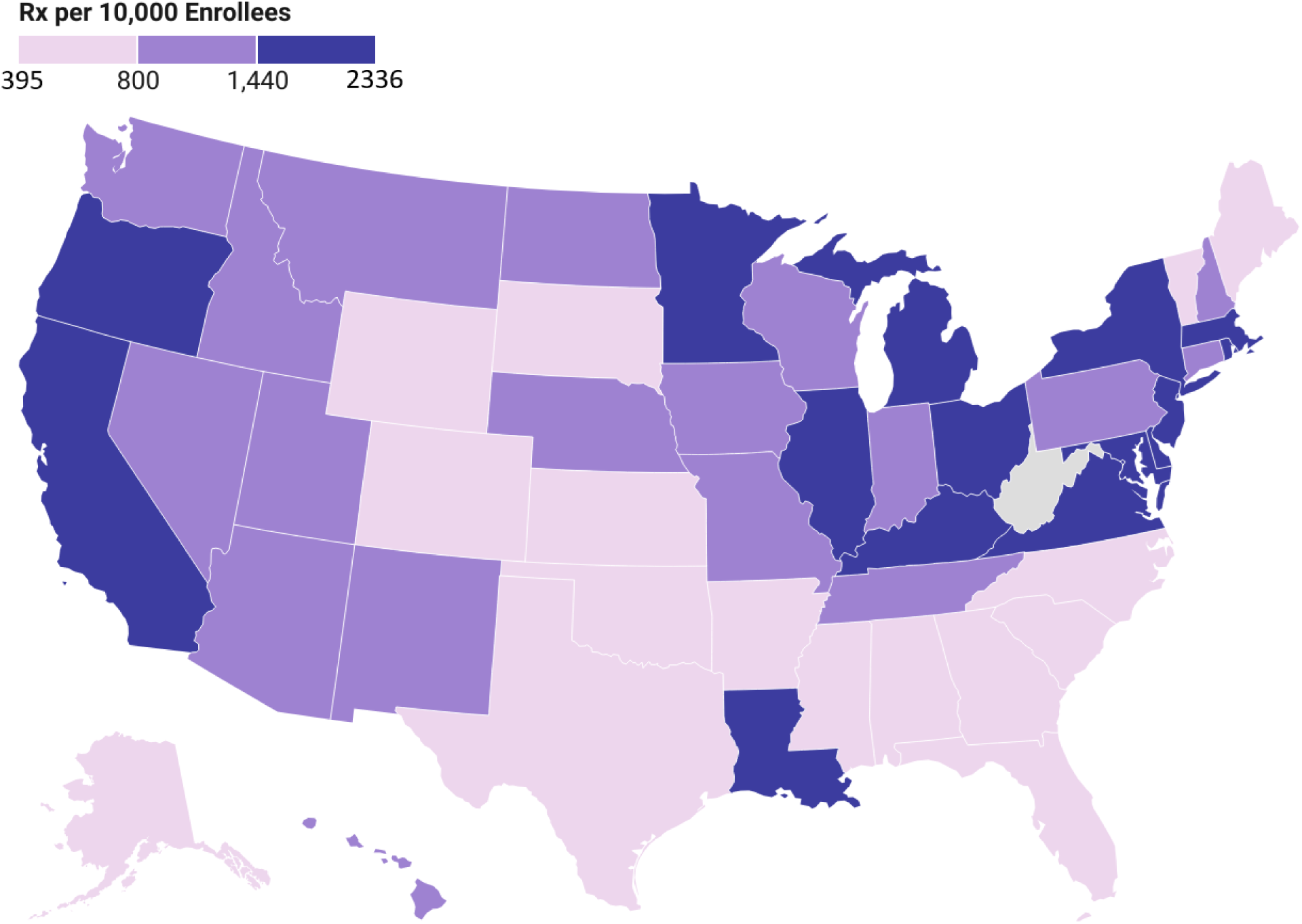
Heatmap of Metformin prescription distributions per 10,000 Medicaid patients in 2020

## Notes

### Author Declarations

The study used ONLY openly available human data from the state utilization data, avaliable on the Medicaid.gov website,

## References

[1] Heart Disease facts [Internet]. Centers for Disease Control and Prevention. Centers for Disease Control and Prevention; 2022 [cited 2022Jul12]. Available from: https://www.cdc.gov/heartdisease/facts.htm#:~:text=About%20659%2C000%20people%20in%20the,1%20in%20every%204%20deaths.

[2] Diabetes [Internet]. Centers for Disease Control and Prevention. Centers for Disease Control and Prevention; 2022 [cited 2022Jul12]. Available from: https://www.cdc.gov/heartdisease/facts.htm#:~:text=About%20659%2C000%20people%20in%20the,1%20in%20every%204%20deaths.

[3] Christou GA, Katsiki N, Blundell J, Fruhbeck G, Kiortsis DN. Semaglutide as a promising Antiobesity Drug. Obesity Reviews [Internet]. 2019 [cited 2022Jun27]; 20(6):805–15. Available from: https://pubmed.ncbi.nlm.nih.gov/30768766/

[4] Blundell J, Finlayson G, Axelsen M, Flint A, Gibbons C, Kvist T, Hjerpsted JB. Effects of once-weekly semaglutide on appetite, energy intake, control of eating, food preference and body weight in subjects with obesity. Diabetes Obes Metab. 2017 Sep;19(9):1242–1251.

[5] Kalra S, Sahay R. A review on semaglutide: An oral glucagon-like peptide 1 receptor agonist in management of type 2 diabetes mellitus [Internet]. Diabetes Therapy. 2020 [cited 2022Jun27];11(9):1965–82. Available from: https://www.ncbi.nlm.nih.gov/pmc/articles/PMC7434819/

[6] FDA approves new drug treatment for Chronic Weight Management, first since 2014 [Internet]. U.S. Food and Drug Administration. FDA; 2021 [cited 2022Jun27]. Available from: https://www.fda.gov/news-events/press-announcements/fda-approves-new-drug-treatment-chronic-weight-management-first-2014

7. “Data.Medicaid.gov.” n.d. Centers for Medicare and Medicaid Services. https://data.medicaid.gov/.

8. Internet Archive. n.d. “Wayback Machine.” Archive.org. https://web.archive.org/.

[9] Scannell, Christopher, John Romley, Rebecca Myerson, Dana Goldman, and Dima M. Qato. 2024. “Prescription Fills for Semaglutide Products by Payment Method.” JAMA Health Forum 5, no. 8 (August): e242026. 10.1001/jamahealthforum.2024.2026.

[10] Berning, Philipp, Rishav Adhikari, Adrian E. Schroer, Yara A. Jelwan, Alexander C. Razavi, Michael J. Blaha, and Omar Dzaye. 2025. “Longitudinal Analysis of Obesity Drug Use and Public Awareness.” JAMA Network Open 8, no. 1 (January): e2457232. 10.1001/jamanetworkopen.2024.57232.

[11] Myers CA, Slack T, Martin CK, Broyles ST, Heymsfield SB. Regional Disparities in obesity prevalence in the United States: A spatial regime analysis [Internet]. Obesity (Silver Spring). 2015 [cited 2022Jul20]; 23(2), 481-487. Available from: https://www.ncbi.nlm.nih.gov/pmc/articles/PMC4310761/.

